# A digital twin methodology using real patient data for sample size reduction in Alzheimer’s disease randomized controlled clinical trials

**DOI:** 10.1101/2025.10.28.25338899

**Authors:** Daniel Andrews, Shirin Golchi, D. Louis Collins, the Alzheimer’s Disease Neuroimaging Initiative

## Abstract

**INTRODUCTION:** Recruitment for Alzheimer’s disease randomized controlled trials (RCTs) is difficult and expensive. To reduce RCT sample sizes, our Digital Twin Trial (DTT) methodology combines an interpretable cognitive decline prediction model with prediction-powered inference.

**METHODS:** For DTT participants, our model identifies similar individuals (“Digital Twins”) from a retrospective database and uses their cognitive scores to predict decline. Predictions adjust observed scores, reducing variance within treatment groups. We simulated 18-month DTTs and standard RCTs using mixed effects models of decline in Alzheimer’s Disease Neuroimaging Initiative subjects meeting lecanemab’s Phase 3 inclusion criteria.

**RESULTS:** Predicted and observed change in Clinical Dementia Rating Sum-of-Boxes correlated at *r* = 0.4. DTTs required 1,855 subjects versus 2,170 for standard RCTs to detect a simulated 25% decline-slowing drug effect at 0.9 power. DTT Type 1 error was consistent with 0.05.

**DISCUSSION:** DTTs could reduce recruitment and cost burdens. Model interpretability could help clinicians trust individualized prognoses.

## 1. Introduction

Disease-targeted drug treatments for Alzheimer’s disease (AD) are needed to slow cognitive decline in patients and reduce the population incidence of dementia.^1,2^ Over 100 clinical trials of AD-targeted drugs were ongoing as of January 2025, in total requiring tens of thousands of participants.^2^ More treatments are being developed, and all will require testing in randomized controlled clinical trials (RCTs).

Recruitment for AD trials is extremely difficult. Trial planners can expect to randomize fewer than one participant per month per trial site.^3^ Larger cohorts increase statistical power to detect drug effects but increase trial costs and total runtime.^3^ Cohorts should also be as small as possible, given the unconfirmed effects of experimental treatments and the need to limit the number of people receiving placebo. Trial planners must find ways to reduce sample sizes while maintaining high power, while still representing patient diversity and the heterogeneity of AD progression.^4^

To address these issues, in this paper we present a Digital Twin methodology that uses patient data from an existing retrospective study to reduce required sample sizes for RCTs of AD-targeted drugs.^5^

Patient data from external studies have already been used to supplement or replace RCT control groups and increase power.^6–9^ However, average differences in disease progression can exist between the trial’s cohort and the external dataset, despite the same inclusion criteria having been applied, for example due to evolving diagnostic standards in AD or from randomness in recruitment.^4,10–13^ This issue can bias observed treatment effects and increase Type 1 or Type 2 error risk. That risk can be mitigated with nuanced approaches such as Bayesian power prior techniques, where the contribution of external data must be calibrated based on their cohort-level similarity to the trial data.^14,15^ However, such techniques can complicate regulatory compliance.^16^

Recently, individualized predictions of natural disease progression over the RCT duration, or Digital Twins, have been combined with novel data analysis techniques that maintain error control, make treatment effect estimates more precise, increase power, and reduce required sample sizes.^15,17–23^ For example, prognostic covariate adjustment is already being used in real-world AD trials.^24^ Prediction-powered inference (PPI) has been proposed for AD trials after originally being demonstrated for genomics, astronomy, and other scientific data.^17,22,25^ To date, however, clinical trial applications of these techniques have relied on generative machine learning models trained on large historical datasets to predict disease progression.^17,26,27^

In this paper, we expand upon an interpretable Digital Twin prediction model that we originally presented at the 2024 Alzheimer’s Association International Conference.^5^ This model does not require generative machine learning. We empirically show that the model’s prediction accuracy is sufficient to enable power gains and Type 1 error control in PPI for RCTs of drugs designed to slow cognitive decline in AD.

In our model, a trial participant is assigned a set of Digital Twins, defined as the participant’s medically most-similar peers at baseline.^5^ These peers are identified in an external retrospective dataset and satisfy the target trial’s inclusion criteria. The average cognitive decline of a participant’s Digital Twins represents the participant’s predicted decline. We use this model in a recently formalized PPI framework that we compare to a simple RCT analysis.^17^ This comparison will communicate our model’s feasibility and benefits in PPI.

First, we explain how our Digital Twin model could be implemented within PPI in a real trial, which we label a Digital Twin Trial (DTT). Then, we evaluate the accuracy of our model’s cognitive decline predictions in real patients from the Alzheimer’s Disease Neuroimaging Initiative who satisfy inclusion criteria of CLARITY AD, the successful Phase 3 trial of the amyloid-targeting drug lecanemab.^28–30^ Next, in simulation studies constrained by CLARITY AD design parameters, we empirically evaluate DTT and standard RCT power to detect the same drug effect.^31^ We also evaluate Type 1 error probability.^32^ These analyses use realistic computer-generated trial simulations derived from the same patient data used to evaluate our model’s prediction accuracy.

## 2. Methods

### 2.1. Prediction-powered inference for clinical trials

Prediction-powered inference (PPI) for clinical trials can be defined as a data analysis framework with two components.^17^ The first component is a model that predicts each participant’s outcome (e.g., a cognitive test score) under natural disease progression. For each Drug group participant, the prediction represents a counterfactual outcome, or the expected cognitive decline had the drug not been delivered. For Placebo group participants, predictions represent the expected disease progression. This process isolates drug and placebo effects at the participant level.

The second PPI component is an analysis model that first uses each participant’s predicted outcome to adjust their real outcome observed in the trial.^17^ Placebo group observations should align with predictions, but an effective drug should yield substantial differences between observations and predictions. Finally, a treatment effect estimator analyzes the Drug versus Placebo group difference in the prediction-adjusted outcomes.

These two PPI components combine to increase statistical power by reducing the outcome variance in the Drug and Placebo groups compared to unadjusted outcomes.^17^

A variety of such combinations have been proposed for PPI and analogous prognostic analysis techniques.^17,26,27^ This paper presents a combination of our new prediction model with a PPI analysis model recently formalized by Poulet et al.^5,17^ We compare this combination to a simple standard RCT analysis.^5,17^

### 2.2. Overview of the Digital Twin model in an example Digital Twin Trial

We apply the label of Digital Twin Trial (DTT) to any RCT that uses our PPI combination. This paper first explains our DT prediction model methodology and how it could be implemented in a PPI analysis of a real-world DTT. We compare this DTT to a simplified standard RCT (sRCT).

In our prediction model, a participant’s predicted outcome equals the mean outcome observed in the participant’s clinically and demographically most-similar peers identified at baseline (see Fig. 1).^5^ These most-similar peers – the Digital Twins – are identified in an External Cohort pulled from an existing retrospective External Study.^5^ All subjects in the External Cohort satisfy the trial’s inclusion criteria. Note that some existing prognostic adjustment techniques label a participant’s predicted outcome(s) as the Digital Twin, while we label a participant’s most-similar peers as the Digital Twin<u>s</u>.^5,17,26,27^

**Figure 1.**
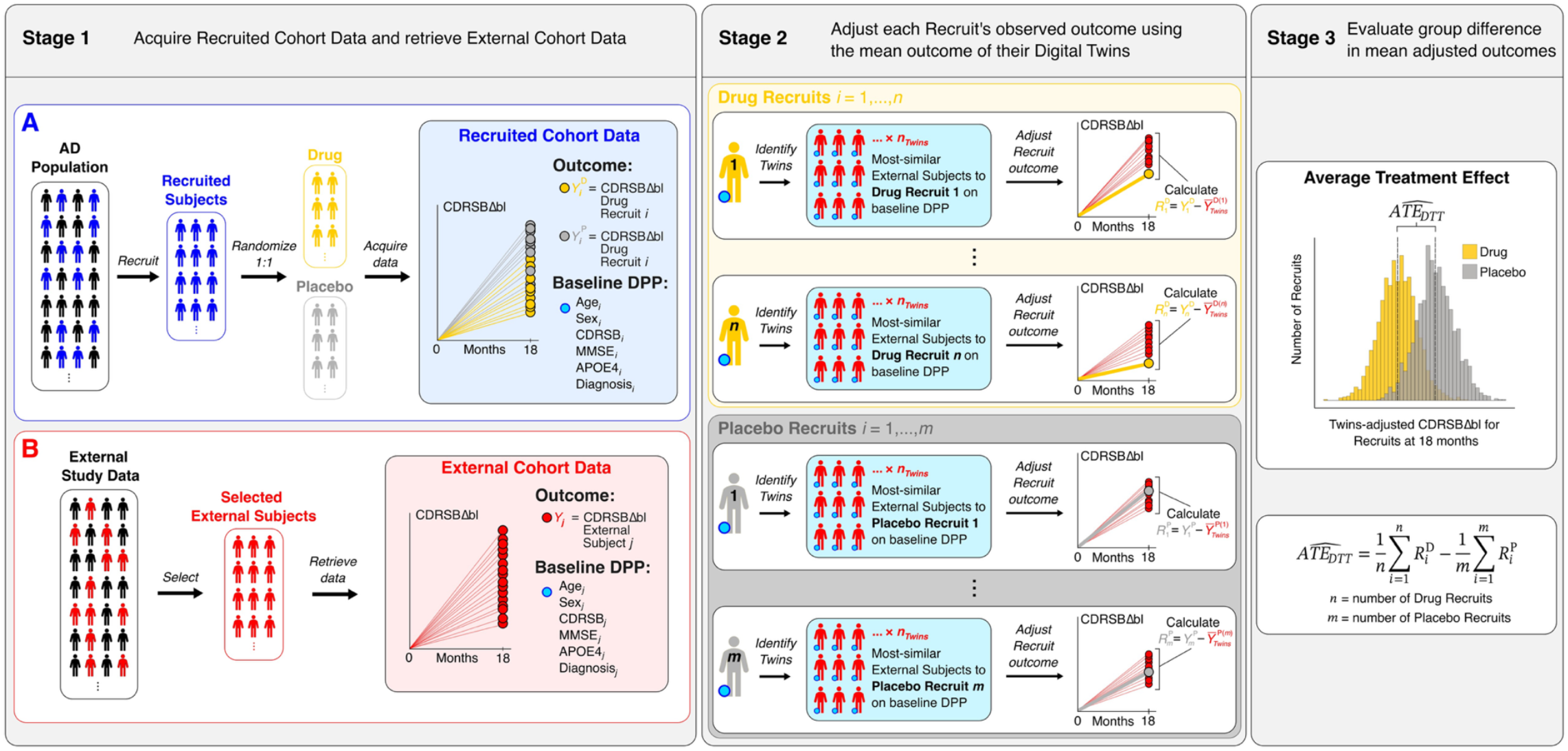
Procedure for a hypothetical 18-month real-world Digital Twin Trial (DTT). <u>Stage 1:</u> A) From the real-world AD population, enroll trial participants as Recruited Subjects and randomize them 1:1 to Drug and Placebo groups. For each Recruit *i* in the Drug or Placebo group, acquire baseline data on age, sex, CDRSB, MMSE, APOE4 status, and diagnosis. The blue dot denotes a Recruit’s baseline data as their Digital Patient Profile (DPP). Next, acquire each Recruit’s primary outcome data, here 18-month CDRSBΔbl, denoted 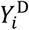 and 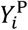 for Recruits *i* in the Drug or Placebo group, respectively. These Recruits comprise the trial’s Recruited Cohort (blue-shaded box in Stage 1-A). B) From an existing retrospective External Study, select the External Subjects who satisfy the trial’s inclusion criteria. Retrieve their baseline DPPs (same variables as above). Also retrieve 18-month CDRSBΔbl data, denoted *Y*_*j*_ for External Subjects *j*. These External Subjects’ data comprise our trial’s External Cohort (red-shaded box in Stage 1-B). <u>Stage 2:</u> For Drug Recruit 1 (topmost white box in the yellow-shaded Drug Recruits box in Stage 2), identify the *n*_*Twins*_ most-similar subjects in the External Cohort at baseline by ranking the similarity of Drug Recruit 1’s baseline DPP to that of each subject in the External Cohort. The most similar *n*_*Twins*_ subjects are Drug Recruit 1’s Digital Twins. Next, adjust the Recruit’s 18-month CDRSBΔbl by subtracting the mean 18-month CDRSBΔbl of their identified Digital Twins. Complete this process for each Drug Recruit *i* = 1, …, *n*, and for each Placebo Recruit i = 1, …, *m* to generate the Twins-adjusted 18-month CDRSBΔbl scores 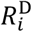 and 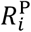, respectively. <u>Stage 3:</u> Use a two-sample *t*-test to evaluate the Drug versus Placebo group difference in mean Twins-adjusted 18-month CDRSBΔbl score. This group difference represents the DTT’s estimated Average Treatment Effect 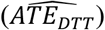.

A real-world DTT could be implemented using a three-stage procedure. Figure 1 describes each stage implemented in a hypothetical 18-month example trial where an AD-targeted drug slows cognitive decline by 25% compared to placebo.

This hypothetical trial replicates key design parameters of the anti-amyloid drug lecanemab’s Phase 3 trial CLARITY AD.^29,30^ This trial is a prime example of a successful modern Phase 3 RCT of a disease-targeted drug for AD.^2^ The example trial arbitrarily includes 1,800 participants, and the primary outcome is the change from baseline to 18 months in the cognitive test score Clinical Dementia Rating Sum-of-Boxes (CDRSBΔbl).^29,30,33^ Sections below describe this DTT and compare it to a simplified standard RCT analysis of the example trial data.

### 2.3. Overview of DTT validation studies

This paper also validates our DTT methodology with three Studies. The combined purpose of these Studies is to show that our proof-of-concept Digital Twin model can predict trial outcomes sufficiently accurately to enable PPI power gains compared to an sRCT while maintaining Type 1 error control. Results will show that our moderately accurate but highly interpretable model enables the benefits of PPI.

In Study 1, we evaluate our model’s prediction accuracy in real patients from the Alzheimer’s Disease Neuroimaging Initiative (ADNI) who satisfy CLARITY AD inclusion criteria.^28–30^ We predict individuals’ 18-month CDRSBΔbl scores. The objective of Study 1 is only to determine the model’s prediction accuracy in one specific cohort, not to demonstrate high generalizable accuracy, nor to compare the accuracy to existing prediction model accuracies.

In Study 2, we use the real patient data from Study 1 to simulate 18-month DTTs and sRCTs.^5,31,34^ The goal here is to empirically evaluate DTT and sRCT power to detect the same cognitive decline-slowing drug effect. This study will let us calculate and compare the required DTT and sRCT sample sizes to reach a statistical power of 0.9.

In Study 3, we use the same simulation procedure as in Study 2 to empirically evaluate DTT and sRCT Type 1 error probability, and to demonstrate Type 1 error control for the DTT using the sRCT probability as the baseline.^32^

### 2.4. Real-world DTT procedure and example

This section explains how a hypothetical real-world DTT could be run and compares it to a simplified sRCT.

#### 2.4.1. Stage 1: Acquire Recruited Cohort data and retrieve External Cohort data

The DTT is a type of RCT that requires (A) a Recruited Cohort, and (B) an External Cohort. The Recruited Cohort is equivalent to that of an sRCT, where subjects are randomized to Drug and Placebo groups. The External Cohort is the subject pool from which Digital Twins will be identified for each Recruited Cohort subject. These cohorts are constructed, respectively, in Parts A and B of our DTT procedure’s Stage 1 (see Fig. 1).

##### 2.4.1.1. Part A: Acquire Recruited Cohort data

In Part A we recruit trial participants from the AD population to form our Recruited Cohort. In our example trial, each Recruit satisfies CLARITY AD’s inclusion criteria for age, abnormal brain amyloid positivity, cognitive diagnostic category of mild cognitive impairment (MCI) or mild AD dementia, Mini Mental State Examination (MMSE) and Clinical Dementia Rating (CDR) scores, and body mass index (BMI).^29,30^

We then acquire baseline data – the Digital Patient Profile (DPP) – for each Recruit. In this example, the DPP includes age, sex, CDRSB, MMSE, apolipoprotein E4 status (APOE4, carrier or not), and diagnosis (MCI or mild AD dementia). In Fig. 1, the Stage 1 Part A panel indicates a Recruit’s DPP with the blue dot.

Next, we randomize the Recruits 1:1 to Drug and Placebo groups and measure the primary outcome, here 18-month CDRSBΔbl. Higher scores indicate worse impairment.^33^ These outcomes are denoted in the Stage 1 panel of Fig. 1 by 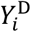 and 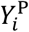 for Recruits *i* in the Drug and Placebo groups, respectively.

##### 2.4.1.2. Part B: Retrieve External Cohort data

In Part B we form our trial’s External Cohort by retrieving the baseline DPPs and the outcome data of External Subjects from an existing retrospective External Study. The outcome and the DPP variables are the same as in Part A. Selected subjects also satisfy the trial inclusion criteria from Part A. For our example, the External Study dataset is the Alzheimer’s Disease Neuroimaging Initiative (ADNI) dataset.^28^ Figure 1 illustrates this process in the Stage 1 Part B panel. In our example, the retrieved outcomes are 18-month CDRSBΔbl scores, denoted in Fig. 1 as *Y*_*j*_ for External Subjects *j*.

#### 2.4.2. Stage 2: Adjust each Recruit’s outcome using their Digital Twins-predicted outcome

In Stage 2 we adjust the observed outcome of each Recruit by subtracting the mean outcome of the Recruit’s Digital Twins. This mean represents the Recruit’s predicted outcome under natural disease progression, accounting for the individualized prognostic factors in the Recruit’s baseline DPP.^5,35^ As described above, the DPP in this example includes age, sex, CDRSB, MMSE, APOE4 status, and cognitive diagnostic category. The Stage 2 panel of Fig. 1 illustrates the procedure. This adjustment is done for all Recruits in both Drug and Placebo groups.

To identify a Recruit’s Digital Twins, we first calculate then rank the similarity of each Recruit’s DPP to that of each External Subject in the External Cohort. We use Gower’s distance to quantify similarity because it automatically scales continuous variables and can accommodate mixed variable types, including numerical, categorical, and binary variables (e.g., continuous age, cognitive diagnostic category, and binary sex).^36^ Smaller Gower distances between DPPs indicate greater similarity, with distances ranging from 0 to 1 (identical to maximally dissimilar.) A Recruit’s top *n*_*Twins*_ most-similar External Subjects are the Recruit’s Twins. For example, an *n*_*Twins*_ = 20 indicates 20 Twins per Recruit.

Equation 1 defines the mean outcome 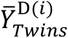 of the Digital Twins of a Recruit *i* in the Drug group. Equation 2 defines that mean for a Recruit *i* in the Placebo group. A Recruit’s Twins are indexed *k* = 1, …, *n*_*Twins*_ Variables 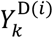 and 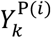 in Eqs. 1 and 2, respectively, denote the outcome of Digital Twin *k* of Recruit *i*.

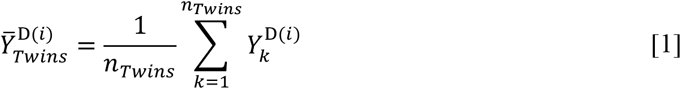

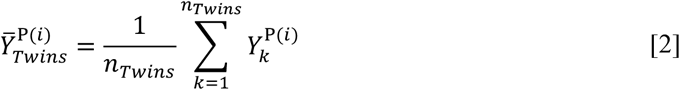

Next, we adjust each Recruit’s observed outcome by calculating its difference from the predicted outcome. Equations 3 and 4 define the resulting Twins-adjusted outcomes as 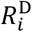 and 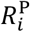.

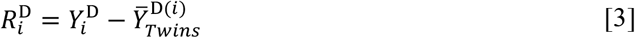

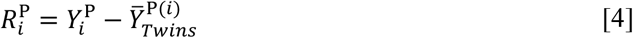

This process assumes that a Recruit’s outcome and those of their Digital Twins are conditionally exchangeable under natural disease progression.^37^ That is, Recruits and Twins with similar prognostic characteristics at baseline are assumed to have natural disease trajectories drawn from the same probability distribution. The Twin outcomes can therefore be used to predict Recruit outcomes in the absence of drug effect.

If this assumption is invalid, bias from the predictions can be injected by this adjustment. However, the adjustment distributes any such bias equally to both treatment groups. A subsequent PPI analysis step cancels out such bias (see section 2.4.3.2).

#### 2.4.3. Stage 3: Evaluate the Average Treatment Effect (ATE)

In Stage 3 we evaluate our trial’s drug effect by estimating an Average Treatment Effect (ATE). We describe the ATE estimator for a simple sRCT analysis of our trial and compare it to our simple DTT estimator.

##### 2.4.3.1. The simple 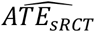 estimator

Equation 5 defines the ATE estimator for our simple sRCT analysis as a group difference in outcome means, and Eq. 6 defines the sampling variance of Eq. 5.^17^ In Eq. 5, the variables *n* and *m* are the numbers of Recruits in the Drug and Placebo groups, respectively. Our example trial includes a drug effect that slows cognitive decline by 25% versus placebo. Assuming adequate power, this effect would be reflected in the Drug group having an estimated mean outcome that is 25% smaller than the Placebo mean.^17^ In this example, statistical inference on Eq. 5 is done via two-sample *t*-test with a two-sided with α = 0.05.

In Eq. 6, 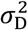 and 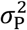 are the population outcome variances under drug and placebo, respectively.^17^

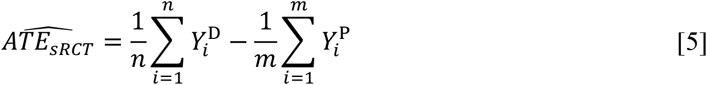

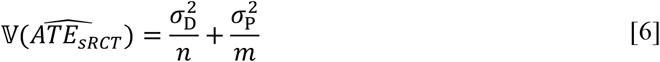

Inference on Eq. 5 answers two questions: (1) On average, did people who received the drug have slower cognitive decline compared to people who received placebo? (2) If so, how much slower? These questions only address the average drug effect on the Drug group’s decline compared to the Placebo group.

##### 2.4.3.2. The simple 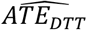 estimator

Equation 7 defines the ATE estimator for our simple DTT analysis as a group difference in mean adjusted outcome, and Eq. 8 defines the sampling variance of Eq. 7.^17^ Assuming adequate power, our posited drug effect would again be reflected in the Drug group having an estimated mean that is 25% smaller than the Placebo mean.^17^ Inference is again done via two-sample *t*-test.

In Eq. 8, 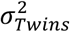 is the population variance of Twins-predicted outcomes, and *ρ*_D_ and *ρ*_P_ are the population correlations between observed outcomes and Twins-predicted outcomes under drug and placebo, respectively.

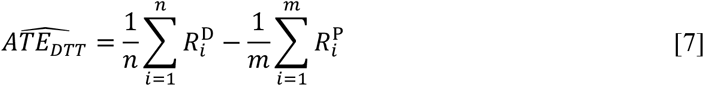

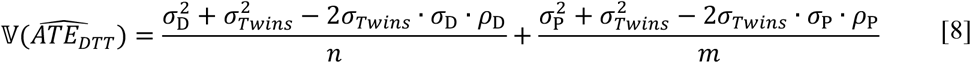

Equation 7 targets a different estimand than Eq. 5, so inference on Eq. 7 answers two questions that are subtly different from those answered by the Eq. 5 inference. The questions answered here are: (1) On average, did people who received the drug decline slower than was expected for them under natural progression compared to people who received placebo? (2) If so, how much slower? These questions address the drug and placebo effect on each person’s individual decline, and the average drug effect on the Drug group’s decline compared to the Placebo group.

The difference in means from Eq. 7 will be the same as for Eq. 5.^17^ However, assuming sufficiently accurate predicted outcomes, the variance of the adjusted outcomes will be lower than the variance of the raw outcomes.^17^ This variance reduction increases the contrast between treatment groups, lowers the standard error of the ATE estimator, and increases statistical power.

By first adjusting the outcomes in both treatment groups, this PPI analysis equally distributes and cancels out any bias injected by the outcome adjustment, which mitigates Type 1 and Type 2 error risk.^17^ This DTT methodology is validated below in three Studies.

### 2.5. Alzheimer’s Disease Neuroimaging Initiative

Data used in the preparation of this article were obtained from the Alzheimer’s Disease Neuroimaging Initiative (ADNI) database (<u>adni.loni.usc.edu</u>). The ADNI was launched in 2003 as a public-private partnership, led by Principal Investigator Michael W. Weiner, MD. The primary goal of ADNI has been to test whether serial magnetic resonance imaging (MRI), positron emission tomography (PET), other biological markers, and clinical and neuropsychological assessment can be combined to measure the progression of MCI and early AD. For up-to-date information, see www.adni-info.org.

### 2.6. Study 1 method: Digital Twin model prediction accuracy and sensitivity analysis

In this Study we evaluate the prediction accuracy of our Digital Twin model using a pool of real AD patients selected from ADNI who satisfy key CLARITY AD inclusion criteria.^28–30^

We split our selected ADNI pool roughly in half along geographical lines. The two resulting cohorts contain subjects from mutually exclusive ADNI collection sites. Subjects from sites ≤ 41 are assigned to a “Recruited Cohort,” and subjects from ADNI sites > 41 are assigned to an “External Cohort.”

This splitting enables out-of-sample prediction: For each subject in the Recruited Cohort, their CDRSBΔbl score at 18 months (within a visit window of approximately ±1 month) will be predicted using their own Digital Twins identified in the External Cohort. This prediction time represents the CLARITY AD duration and the visit window represents a realistic variation in observation times across participants.^29,30^

These observation parameters will also be used in our simulated trials in Studies 2 and 3. The described Recruited and External Cohorts will also be used to generate the correspondingly named cohorts for the simulations in Studies 2 and 3.

#### 2.6.1. Participant characteristics of the Recruited Cohort and the External Cohort

The selected pool of ADNI subjects satisfy baseline clinical criteria for MCI or mild AD dementia.^28–30^ Each subject is positive for abnormal amyloid levels at baseline on a cerebrospinal biomarker (Aβ42 ≤ 980 pg/ml) or a PET biomarker (PiB standardized uptake value ratio [SUVR] > 1.2 or AV45 SUVR > 1.11).^38–40^ Subjects at baseline are between 50 and 90 years old, have MMSE scores ≥ 22 and ≤ 30, are APOE4 genotyped, and have a BMI > 17 and < 35. Subjects with MCI have a baseline global CDR score of 0.5 and a CDR Memory Box ≥ 0.5. Subjects with mild AD dementia have a baseline global CDR of 0.5 or 1.0, and a CDR Memory Box ≥ 0.5. Of these subjects, we select only those who have at least two observations of CDRSBΔbl and at least 1.6 years total follow up time.

As described in section 2.6, the resulting subject pool is split into a Recruited Cohort and an External Cohort.

#### 2.6.2. Modeling and interpolation of 18-month CDRSBΔbl scores

Most of the selected ADNI subjects do not have CDRSBΔbl scores available specifically within an approximate ± 1 month window around 18 months. To address this issue, we use a linear mixed effect model to map subjects’ CDRSBΔbl trajectories and then interpolate a CDRSBΔbl score for each subject at a random timepoint in the 18 ± 1 month window. Interpolated scores are uniformly distributed in this window. For simplicity in the explanations below, we label all interpolated scores as “18-month CDRSBΔbl” scores, despite scores being interpolated in the described window.

One model is fitted and used for interpolation in the Recruited Cohort, and, separately, another model is fitted and used in the External Cohort. Observations up to 60 months are included in the training data. We fit the Eq. 9 model in *R* with restricted maximum likelihood using the function *lmer* in package *lme4*.^41,42^

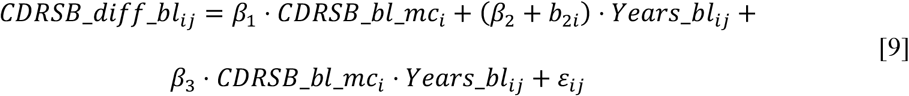

The outcome variable *CDRSB*_*diff*_*bl*_*ij*_ represents CDRSBΔbl for subject *i* at observation *j*. The fixed effect variable *CDRSB*_*bl*_*mc*_*i*_ represents a subject’s median-centered CDRSB score at baseline. Median centering makes parameter estimates more robust. The term *Years*_*bl*_*ij*_ represents a subject’s number of years elapsed since baseline for observation *j*. The *β*_*k*_ terms are the fixed effect coefficients.

Random slopes are encoded by subject to obtain estimates of subject-specific slope deviations *b*_2*i*_ from the cohort-level slope *β*_2_. This slope information is required to interpolate an 18-month CDRSBΔbl for each individual subject. No intercepts are encoded because CDRSBΔbl at baseline is zero for all subjects. The *ε*_*ij*_ term represents the residuals.

Each Recruited Cohort subject’s 18-month CDRSBΔbl is interpolated by evaluating Eq. 9 using the fitted cohort model’s estimated parameters, the subject’s observed *CDRSB*_*bl*_*mc*_*i*_, and *Years*_*bl* = 1.5 + *δ*_*i*_. The term *δ*_*i*_ of subject *i* is a random value that establishes a deviation in observation time from 18 months, for visit windowing. Interpolated CDRSBΔbl scores are rounded to the standard 0.5 increments.^33^ The same procedure is applied to subjects in the External Cohort.

#### 2.6.3. Digital Twin prediction of 18-month CDRSBΔbl scores

Prediction is done as described in section 2.4.2. The “observed” 18-month CDRSBΔbl score of each subject in the Recruited Cohort is predicted using the mean “observed” 18-month score of their most-similar baseline Digital Twins identified in the External Cohort.

In both Cohorts, each subject’s observed score is defined as the value interpolated inside the approximately ± 1 month window around 18 months, as described in 2.6.2. The visit windowing in the External Cohort simulates the realistic trial scenario of the External Cohort having a similar visit window to that of the trial’s Recruited Cohort.

Baseline DPP similarity is quantified by Gower’s distance. No cross validation is done because of the limited number of selected ADNI subjects and our splitting of the subject pool by ADNI site.

We evaluate the Pearson correlation (*r*) between predicted and observed 18-month CDRSBΔbl scores for different combinations of Digital Twin identification parameters. Specifically, we evaluate the described correlation at each of 39 different *n*_*Twins*_ values in the range 2 to 40 for three different sets of prognostic DPP variables (described below). The given DPP variables were selected because they are widely used in the prognosis of AD and are available on most subjects the ADNI database.

Our goal here is to identify which DPP and which *n*_*Twins*_ value yield the highest Pearson *r* between predicted and observed CDRSBΔbl scores. The identified DPP and *n*_*Twins*_ are used in Studies 2 and 3.

The first DPP is an interpretable and balanced set of core demographic, cognitive, and diagnostic measures at baseline known to be associated with AD cognitive decline: age, sex, APOE4 status (1 = carrier, 0 = non-carrier), mild AD dementia status (1 = positive, 0 = negative), early MCI status, late MCI status, CDRSB, and MMSE.^35^

The second DPP contains a minimal set of prognostic baseline variables: age, sex, APOE4 status, and CDRSB. This set will let us evaluate the added predictive value of including the additional variables in DPP sets 1 and 3.

The third DPP is a maximal set that contains all variables from the first DPP as well as baseline BMI (a metabolic health indicator associated with AD cognitive decline) and number of years of education (a proxy for cognitive reserve).^43,44^ This set will let us evaluate whether these general health and lifestyle factors add predictive value compared to the DPP set 1.

Finally, we evaluate the described Pearson *r* when a subject’s Twins are selected at random.

### 2.7. Study 2 method: Standard RCT and DTT power analysis

The goal of this Study is to answer the question: How many Recruits are required for our DTT analysis and an sRCT analysis to detect the same 25% cognitive decline-slowing drug effect versus placebo over 18 months, with power = 0.9 and a two-sided α = 0.05? We use DTT and sRCT simulations to answer this question empirically.

All simulations use the Recruited Cohort and External Cohort data from Study 1 and are constrained by inclusion criteria and the 18-month duration of CLARITY AD.^29^ The question above is answered using our recently proposed trial simulation and power analysis procedure.^31,34^

First, we simulate one set of 3,000 DTTs and one set of 3,000 sRCTs for each of nine sample size values from 1,500 to 2,300 in 100 subject increments. For a given DTT or sRCT sample size, the empirical power is the proportion of 3,000 simulations with a statistically significant ATE.

For the sRCT, Eq. 5 defines the ATE estimator. For the DTT, the estimator is Eq. 7. Inference on both is done via two-sample *t*-test with a two-sided with α = 0.05.

The DTT sample size required for 0.9 power is the smallest sample size that reaches an empirical power of 0.9. The same statement applies to the sRCT. We now describe how we apply this procedure to the sRCT, then to the DTT.

#### 2.7.1. Standard RCT simulation and power analysis procedure

##### 2.7.1.1. Stage 1: Create the baseline Simulated Recruited Cohort

We start by simulating an sRCT that has a Simulated Recruited Cohort containing 1,500 subjects, i.e., the first value in our 1,500 to 2,300 range. The Recruited Cohort dataset from Study 1 has fewer than 1,500 subjects, so here we sample subjects from that cohort with replacement to obtain 1,500 for our Simulated Recruited Cohort. This sampling also maintains a realistic 3:2 ratio for MCI to mild AD subjects, approximating the ratio of the real CLARITY AD trial.^29,30^

We use the same dataset structure for our DTT and sRCT simulations for consistency across Studies. Each sampled subject is thus represented by a vector that includes (1) the subject’s baseline DPP from section 2.6.3, and (2) the subject’s cognitive decline slope parameter from the Eq. 9 model fit to the Recruited Cohort from Study 1. The resulting dataset is our baseline Simulated Recruited Cohort data.

##### 2.7.1.2. Stage 2: Simulate outcomes and evaluate the 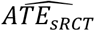

Here we use our trial simulation technique to simulate 18-month CDRSBΔbl scores for the sRCT.^5,31,34^ This process generates our Simulated Recruited Cohort’s outcome data.

First, we randomize subjects in our Simulated Recruited Cohort 1:1 to Drug and Placebo groups. Next, we interpolate each subject’s 18-month CDRSBΔbl score using the method from Study 1. A window of ±1 month is also added around 18-months to represent realistic variation in observation time between subjects. We also inject the drug effect as a 25% reduction of Drug group subjects’ linear CDRSBΔbl slope compared to the Placebo CDRSBΔbl slope. The Placebo slope here is equal to the estimated coefficient *β*_2_ from the fitted Eq. 9 model. To simulate between-subject variation in drug effect, random noise is added to the drug effect of each Drug group subject.

Noise is also added to each model parameter prior to interpolation and to each interpolated CDRSBΔbl score. This process accounts for model uncertainty and simulates measurement error. We round the interpolated CDRSBΔbl scores to the standard 0.5 increments.^33^

Finally, we apply a two-sided two-sample *t*-test with α = 0.05 to our simulated Drug and Placebo groups’ 18-month CDRSBΔbl scores. This step performs inference on the Eq. 5 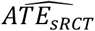 estimator, where the drug effect is the group difference in mean 18-month CDRSBΔbl.

##### 2.7.1.3. Stage 3: Repeat simulations and calculate power

We repeat the above sRCT simulation procedure 3,000 times and calculate the empirical power to detect our injected drug effect. Each simulation has the same duration and 1,500-subject sample size. However, due to the random sampling described above, each simulation will have a unique set of cognitive decline trajectories and therefore a unique observed 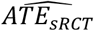. We repeat the above power calculation for the remaining sample sizes from 1,600 to 2,300 and plot the power curve. From this curve we visually identify the approximate sample size value that satisfies our 0.9 power criterion.

#### 2.7.2. DTT simulation and power analysis procedure

The procedure to simulate one DTT overlaps substantially with the sRCT simulation. However, the DTT simulation follows the stages in Fig. 1 but includes an additional stage for the power analysis.

##### 2.7.2.1. Stage 1: Create the Simulated Recruited Cohort and retrieve the External Cohort

Here we generate baseline and outcome data for a Simulated Recruited Cohort with 1,500 subjects following the same steps as in sections 2.7.1.1–2. The resulting dataset thus includes Drug and Placebo group Recruits, each with their baseline DPP and an interpolated 18-month CDRSBΔbl score (see Fig. 1 Stage 1-A).

The External Cohort here is equivalent to that in Study 1, including the given baseline DPP and interpolated 18-month CDRSBΔbl scores for all subjects in the cohort (see Fig. 1 Stage 1-B).

##### 2.7.2.2. Stage 2: Adjust each Recruit’s outcome using the Digital Twins predicted outcome

In this stage we identify the set of Digital Twins of each Recruit in our Simulated Recruited Cohort using the process described in section 2.6.3. The Twins are identified in the External Cohort. The mean 18-month CDRSBΔbl score from a Recruit’s Twins represents the Recruit’s predicted 18-month CDRSBΔbl.

The DPP variable set and *n*_*Twins*_ value used in the Twin identification step of this study are the ones that yielded the highest Pearson *r* between observed and predicted 18-month CDRSBΔbl scores in Study 1 (see section 2.6.3).

Each Recruit’s predicted outcome is subtracted from the Recruit’s observed outcome to produce the Twins-adjusted outcome. This adjustment is done for each Recruit in the Drug and Placebo groups, as described in 2.4.2 (see Fig. 1 Stage 2).

##### 2.7.2.3. Stage 3: Evaluate the 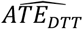

Here we apply a two-sided two-sample *t*-test with α = 0.05 to perform inference on the Eq. 7 estimator. The 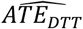 here is the group difference in mean simulated Twins-adjusted 18-month CDRSBΔbl scores (see Fig. 1 Stage 3).

##### 2.7.2.4. Stage 4: Repeat DTT simulations and calculate power

This stage is required only for power analysis. As in our sRCT analysis, here we run 3,000 DTT simulations and calculate power for the 1,500-subject sample size. Each simulation will have a unique set of decline trajectories and 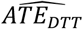. We repeat the power calculation for the remaining sample sizes from 1,600 to 2,300, plot the power curve, and visually identify the smallest sample size that satisfies our 0.9 power criterion.

### 2.8. Study 3 method: Standard RCT and DTT Type 1 error probability analysis

In this Study we use 3,000 trial simulations to empirically evaluate the Type 1 error probability of our sRCT and our DTT for each sample size value from 1,500 to 2,300 separated by 100 subject increments.

We run these simulations as in Study 2, but here we do not inject any drug effect.^32^ However, we still randomize simulated Recruits to Drug and Placebo groups. Therefore, in the inference steps, a statistically significant ATE indicates a Type 1 error. In this scenario, the proportion of simulations with a statistically significant ATE represents the empirical Type 1 error probability.

This Type 1 error analysis generates a Type 1 error probability versus sample size curve for our sRCT and for our DTT. We consider Type 1 error risk controlled in our DTT if the probability at each sample size is not significantly different from 0.05, nor from the corresponding sRCT probability. Significance in this step is evaluated with a chi-squared test with α = 0.05.

## 3. Results

### 3.1. Study 1 results: Digital Twin model prediction accuracy and sensitivity analysis

#### 3.1.1. Participant demographics and observation statistics

From the cohort of available ADNI subjects, 520 satisfied our CLARITY AD and observation scheme inclusion criteria. This subject pool was split into (A) a Recruited Cohort (n = 261) that included subjects from ADNI sites ≤ 41, and (B) an External Cohort (n = 259) including subjects from sites > 41. All subjects had at least two CDRSBΔbl observations. Table 1-A and B provide key demographic characteristics of each cohort, respectively.

**Table 1.** Demographics of selected ADNI subjects included in (A) the Recruited Cohort and (B) the External Cohort used in Studies 1–3. The *p*-values correspond to estimated differences for the given metrics between the two cohorts.

| | **A) Recruited**<br>**Cohort**<br>n = 261 from<br>ADNI sites $\leq 41$ | **B) External**<br>**Cohort**<br>n = 259 from<br>ADNI sites $> 41$ | ***p*-value** |
| --- | --- | --- | --- |
| Age (mean years<br>± std. deviation) | 73.05 ± 7.20 | 74.00 ± 6.95 | 0.1233 |
| Education (mean<br>years ± std.<br>deviation) | 16.08 ± 2.78 | 15.67 ± 2.89 | 0.1014 |
| Female subjects | 102 | 104 | 0.8023 |
| APOE4 carriers | 172 | 177 | 0.5538 |
| Mild AD<br>dementia | 39 | 53 | 0.0990 |
| Mild cognitive<br>impairment | 222 | 206 | 0.0990 |

Recruited Cohort subjects had a mean total follow up time of 3.41 **±** 0.96 years. Value ranges here indicate standard deviations. The minimum and maximum follow up times across subjects were 1.74 years and 5.00 years, respectively. The mean number of CDRSBΔbl observations per subject was 5.40 **±** 1.30. This cohort included 102 female and 159 male subjects. The mean baseline age was 73.05 ± 7.20 years and the mean number of years of education was 16.08 ± 2.78. 172 subjects were APOE4 carriers. At baseline, 39 subjects had mild AD dementia and 222 had MCI.

External Cohort subjects had a mean total follow up time of 3.29 **±** 1.02 years. The minimum and maximum follow up times across subjects were 1.90 years and 5.00 years, respectively. The mean number of CDRSBΔbl observations per subject was 5.20 **±** 1.33. This cohort had 104 female and 155 male subjects. The mean baseline age was 74.00 ± 6.95 years, and the mean years of education was 15.67 ± 2.89. 177 subjects were APOE4 carriers. At baseline, 53 subjects had mild AD dementia and 206 had MCI.

There were no statistically significant differences between the cohorts on continuous metrics (two-sided two-sample *t*-test with α = 0.05) or count metrics (two-sample chi-squared test with α = 0.05). Table 1 provides the associated *p*-values.

#### 3.1.2. Modeling results

Table 2 provides parameter estimates from the Eq. 9 model fit to (A) the Recruited Cohort data and (B) the External Cohort data of Studies 1–3. The linear slopes of the two models were not significantly different, given the *p*-value = 0.24073 at a two-sided α = 0.05 for a *z*-test on the difference in *β* values for the “Years elapsed since baseline” terms. The difference in *β* values for the interaction term was also not significantly different (*p*-value = 0.74459), but the *β* difference for median-centred baseline CDRSB was significantly different (*p*-value = 0.00415). The Recruited and External Cohort Models had Nakagawa conditional *R*^2^ values of 0.857 and 0.889, respectively.^45,46^ These *R*^2^ values indicate that these models adequately map cohort-level and subject-level cognitive decline trajectories.

**Table 2.**
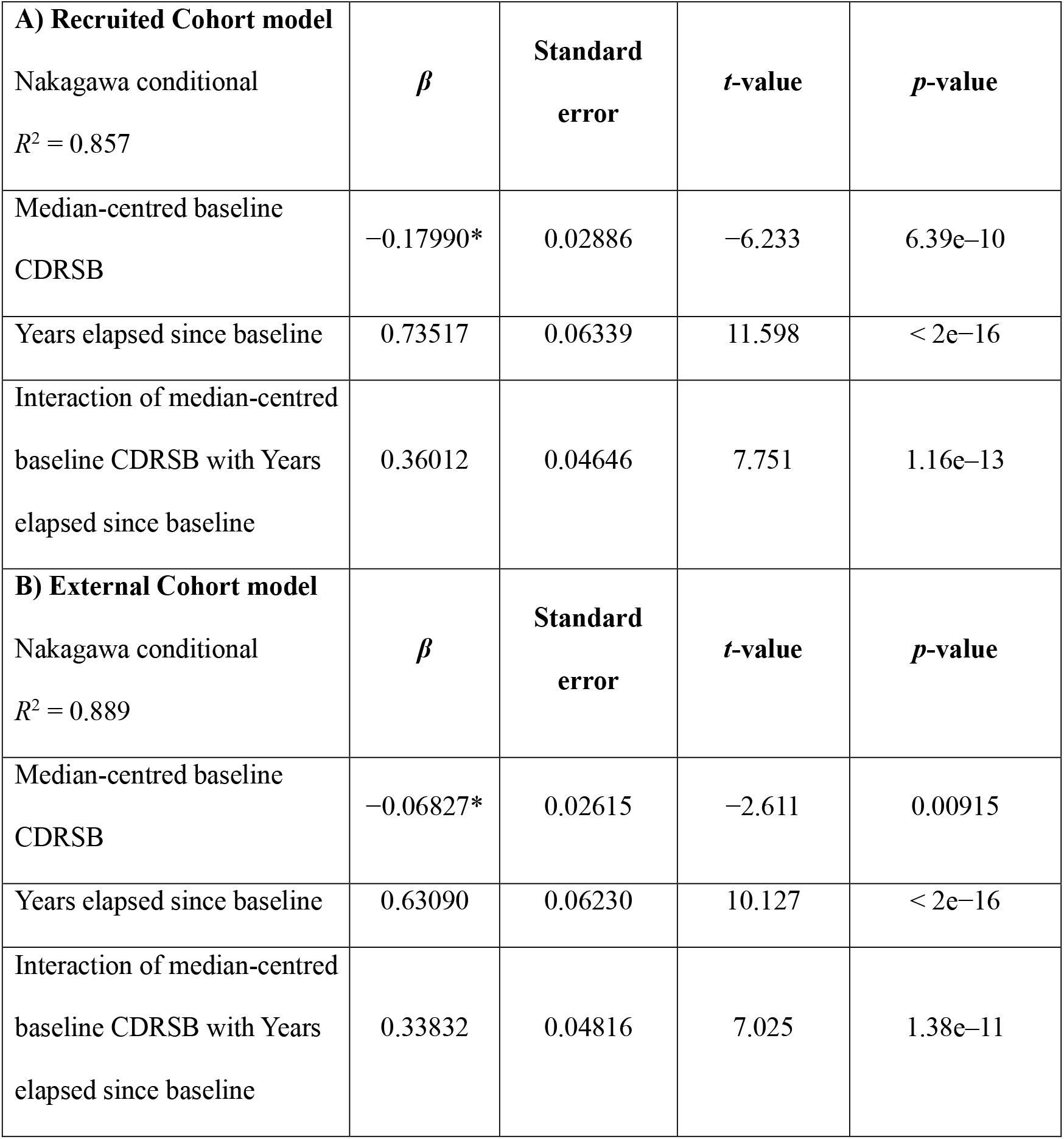
Parameter estimates from the Eq. 9 linear mixed effects model fit to (A) the Recruited Cohort data and (B) the External Cohort data of Studies 1–3. A star on a coefficient estimate indicates a statistically significant difference (*p*-value < 0.05) between the two models.

Figure 2-A shows CDRSBΔbl trajectories over time of individual subjects in the Recruited Cohort as well as the Eq. 9 model trend. Figure 2-B shows CDRSBΔbl data for selection of nine subjects from this cohort, with subject-specific trend lines indicated. These individualized trends represent the linear trends from the Eq. 9 model equation with its estimated cohort-level and subject-specific parameters, and the subject’s own baseline median-centred CDRSB score. In Study 1, a subject’s interpolated CDRSBΔbl score at a specific timepoint within the ± 1 month window around 18 months thus corresponds approximately to a CDRSBΔbl value on the subject’s own trend line, but rounded to the nearest 0.5 value. Figure 2-C and D show the corresponding plots for subjects in the External Cohort.

**Figure 2.**
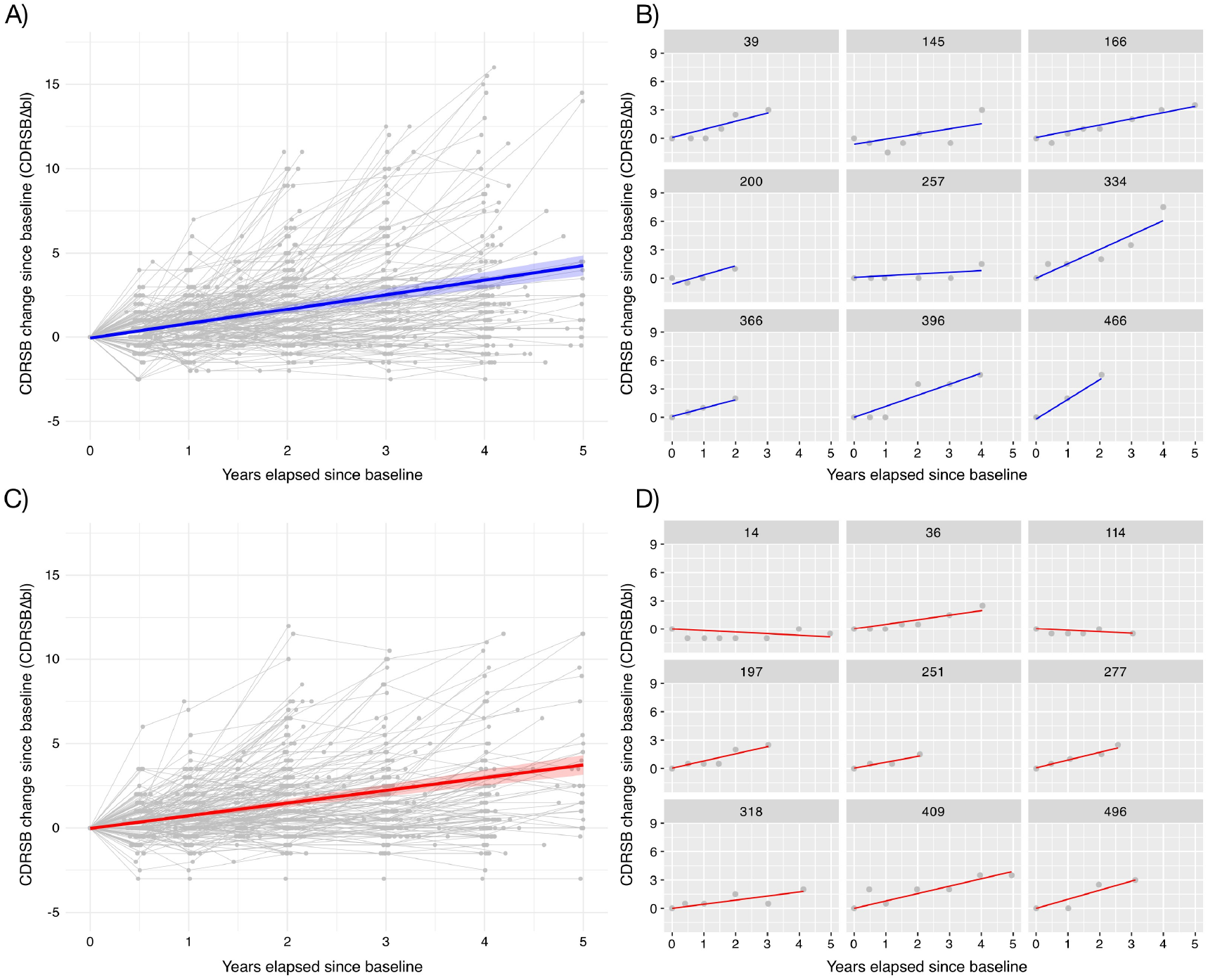
(A) CDRSBΔbl scores over time in real subjects from the Recruited Cohort of Studies 1–3. Subject-specific CDRSBΔbl trajectories are indicated in grey. The blue line and confidence interval indicates the Eq. 9 model trend fit to this cohort. (B) CDRSBΔbl trajectories in nine individual subjects from the Recruited Cohort. Subjects are identified by their index in the original pool of 520 ADNI subjects. Each blue line indicates a subject-specific linear trend derived using the subject’s own median-centred baseline CDRSB score and the required cohort-level and subject-specific estimated parameters from the Recruited Cohort’s Eq. 9 model. (C) CDRSBΔbl scores in subjects from the External Cohort of Studies 1–3. The red line indicates the Eq. 9 model trend fit to this cohort. (D) CDRSBΔbl scores over time in nine subjects from the External Cohort. Subjects are identified by their index in the original pool of 520 ADNI subjects. Red lines indicate the subject-specific trends constructed using the required estimated parameters from the External Cohort’s Eq. 9 model.

#### 3.1.3. Accuracy of Digital Twin 18-month CDRSBΔbl predictions

Figure 3 shows the Pearson *r* of observed and predicted 18-month CDRSBΔbl scores for *n*_*Twins*_ values from 2 to 40. The error bars represent Fisher’s *z* confidence intervals.^47^

**Figure 3.**
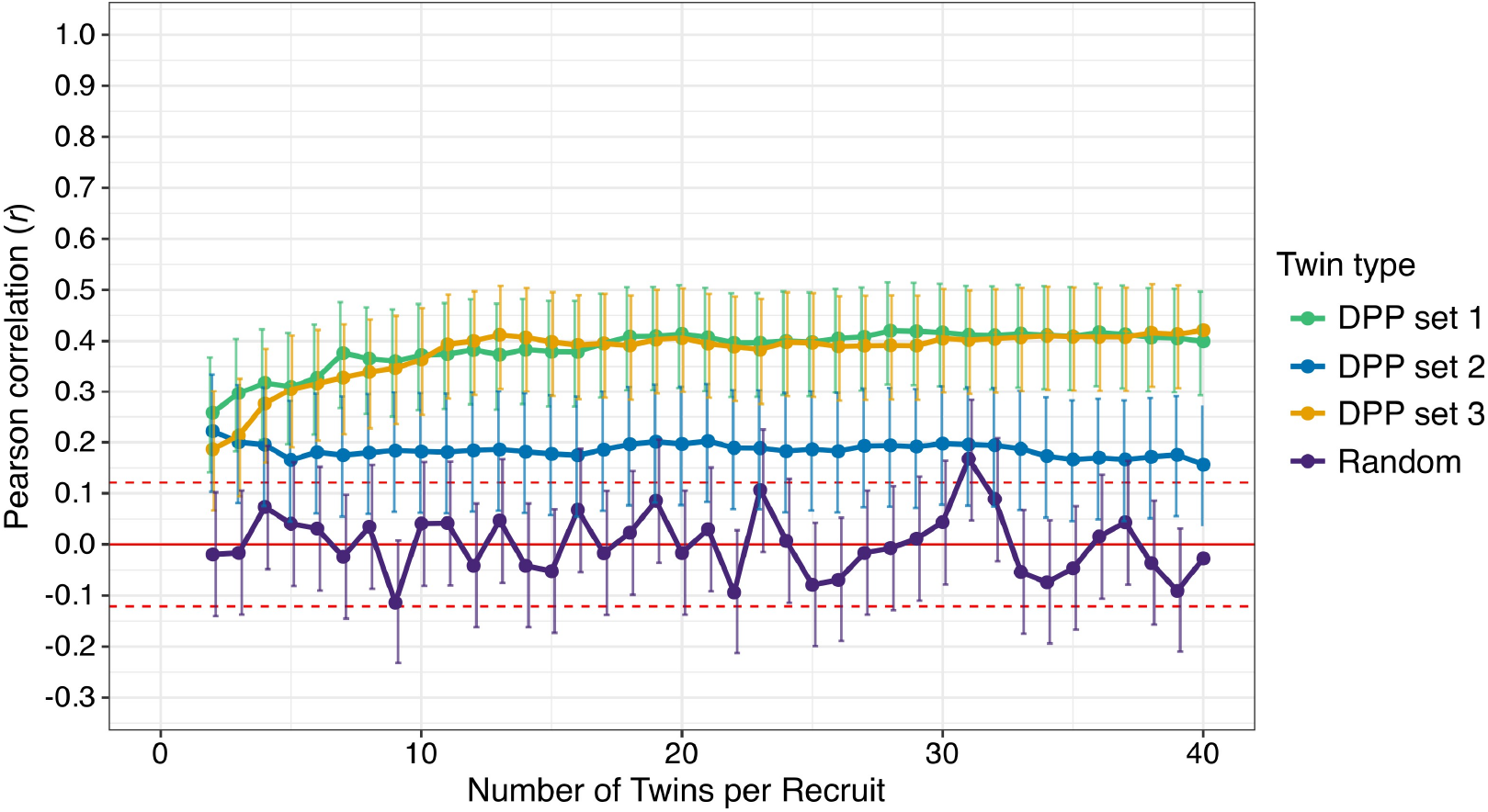
Pearson *r* of observed and predicted 18-month CDRSBΔbl scores across a range of *n*_*Twins*_ values from 2 to 40. Dots indicate estimated *r* values with Fisher’s *z* confidence intervals with a two-sided α = 0.05. The Twin type colours indicate which set of baseline Digital Patient Profile (DPP) variables were used in the Twin identification step. Twins type Random implies that Twins were selected at random for each Recruit. DPP set 1 includes baseline age, sex, APOE4 carrier status, cognitive diagnostic category (AD dementia, early MCI, or late MCI), CDRSB, and MMSE. DPP set 2 includes baseline age, sex, APOE4 carrier status, and CDRSB. DPP set 3 includes all variables in DPP set 1 as well as number of years of education and baseline BMI.

The *r* values for DPP sets 1 and 3 reached an asymptote of approximately 0.4 at *n*_*Twins*_ = 20. Across *n*_*Twins*_ values, the DPP set 1 containing core clinical and demographic prognostic variables yielded correlations not significantly different from those of the maximal DPP set 3, as the *r* values for these DPPs fell inside the opposing DPP’s *r* confidence intervals. The inclusion of BMI and education in DPP 3 thus did not substantially improve prediction accuracy over DPP 1.

The minimal DPP set 2 yielded worse prediction accuracies than DPP sets 1 and 3 for *n*_*Twins*_ > 2, highlighting the added predictive value of a DPP that also includes baseline cognitive diagnostic category and MMSE. The DPP set 2 also yielded worsening prediction accuracy as *n*_*Twins*_ was increased beyond 21.

For predictions made using randomly selected Twins, *r* values track the zero line, with only one value falling outside the zero line confidence interval. All *r* values for DPP sets 1 and 3 lie outside the intervals of corresponding *r* values from the Random curve, and vice versa.

Based on these results, for Studies 2 and 3 we use DPP set 1 with *n*_*Twins*_ = 20. This combination balances parsimony and prediction accuracy. This DPP and *n*_*Twins*_ combination gives a correlation of 0.414, an out-of-sample *R*^2^ of 0.162, a mean absolute error of 0.829, and a root-mean-squared error of 1.094 (Note: CDRSBΔbl test score range is –18 to 18).^33^

### 3.2. Study 2 results: Standard RCT and DTT power analysis

Figure 4 shows sRCT and DTT the empirical power values for each of the 9 sample sizes from 1,500 to 2,300 Recruits. Error bars indicate Wilson confidence intervals for binomial proportions with a two-sided α = 0.05.^48^ The solid red line indicates the 0.9 power threshold and the dashed red lines indicate the confidence interval.

**Figure 4.**
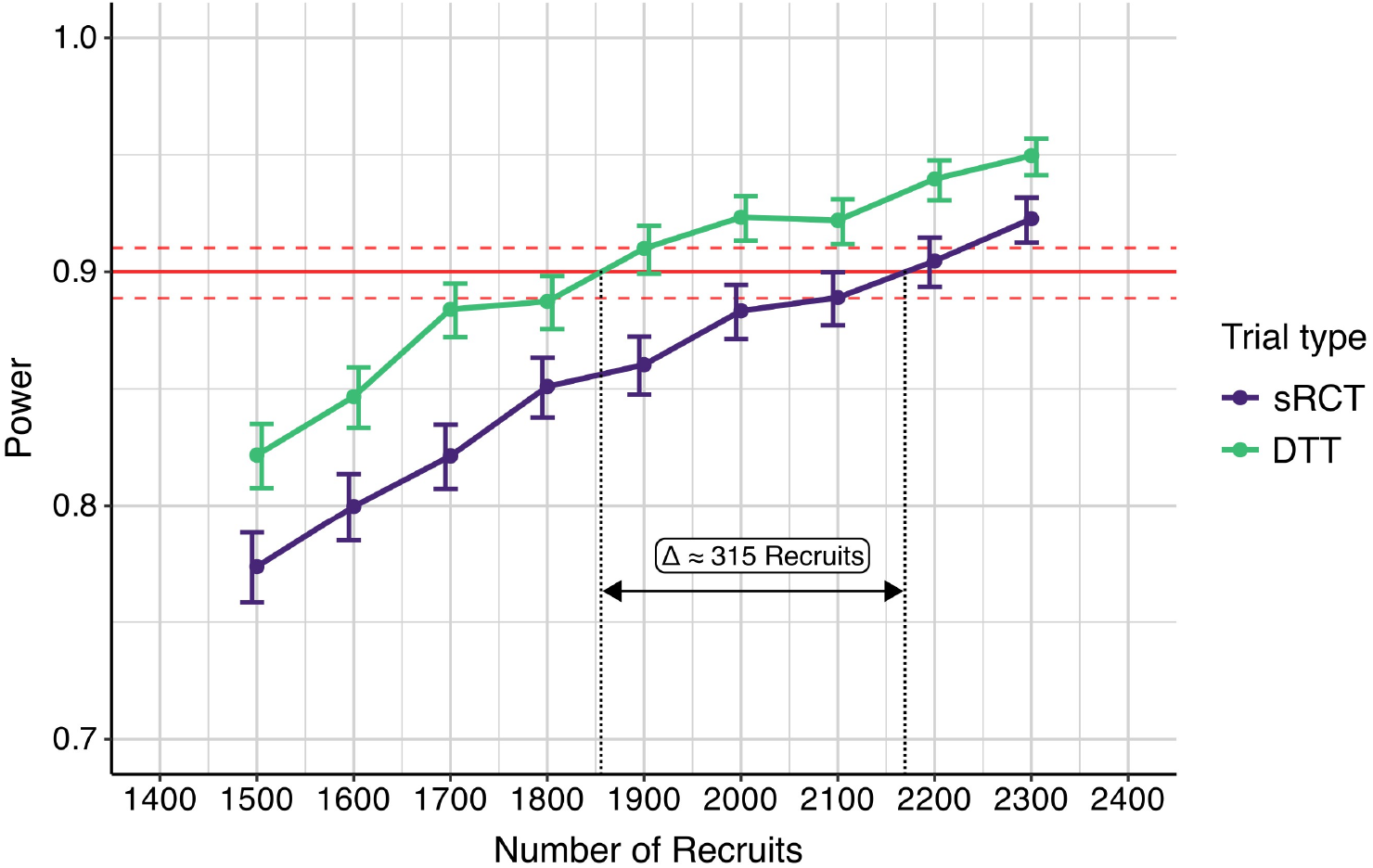
Empirical power curves for the standard RCT (sRCT) and the Digital Twin Trial (DTT). Power was calculated in discrete sets of 3,000 trial simulations at each of 9 sample size values (number of Recruits) from 1,500 to 2,300 in steps of 100. Each power value has its Wilson confidence interval for binomial proportions for a two-sided α = 0.05. The solid red line indicates 0.9 power, and the red dashed lines indicate the confidence interval. The DTT reaches 0.9 power at approximately 1,855 Recruits, while the sRCT reaches 0.9 power at approximately 2,170 Recruits.

Figure 4 indicates that the DTT requires approximately 1,855 Recruits to reach 0.9 power while the sRCT requires approximately 2,170 Recruits. The DTT thus required approximately 315 (14.5%) fewer Recruits than the sRCT. DTT power was 0.042 higher than sRCT power on average across sample sizes. The power curves are not smooth because power values were calculated within discrete sets of 3,000 simulations at each sample size.

### 3.3. Study 3 results: Standard RCT and DTT Type 1 error probability analysis

Figure 5 shows sRCT and DTT Type 1 error probability for each of the 9 sample size values from 1,500 to 2,300 Recruits, with Wilson confidence intervals for a two-sided α = 0.05.^48^ The solid and dashed red lines indicates the 0.05 probability threshold and its confidence interval, respectively.

**Figure 5.**
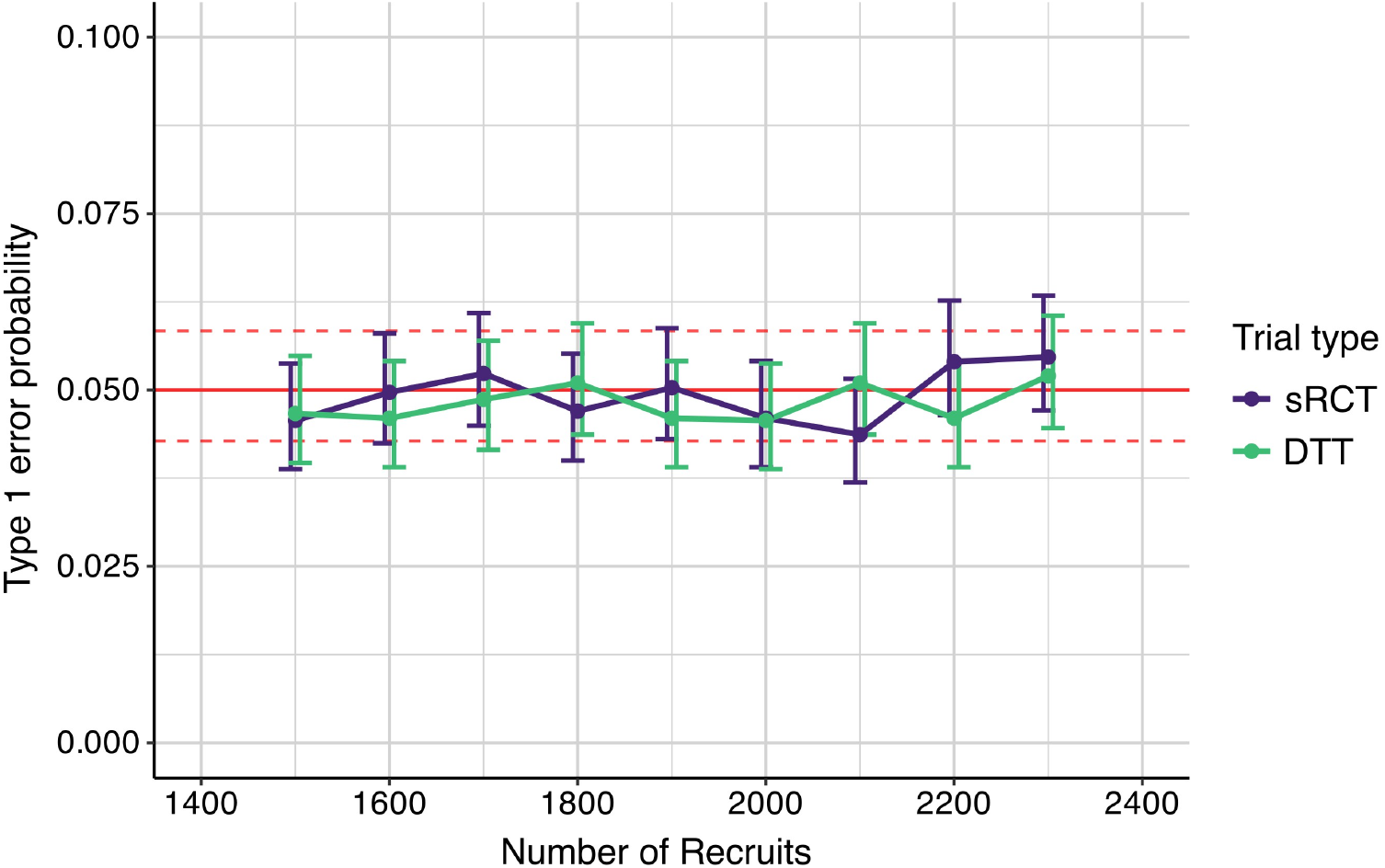
Empirical Type 1 error probability curves for the standard RCT (sRCT) and the Digital Twin Trial (DTT). Probability was calculated in discrete sets of 3,000 trial simulations at each of 9 sample size values from 1,500 to 2,300 in steps of 100. Error bars indicate Wilson confidence intervals. The red lines indicate 0.05 probability and its confidence interval.

There was no statistically significant difference in probability between the DTT and sRCT at each sample size in a two-sided chi-squared test with α = 0.05. No estimated probability value was significantly different from 0.05 using the described chi-squared test. The curves are not smooth because probabilities were calculated within discrete sets of 3,000 simulations at each sample size.

## 4. Discussion

This paper presents an interpretable Digital Twin prediction model of cognitive decline and combines it with prediction-powered inference for clinical trials.^5,17^ We explain how this combination could be implemented in a real-world Digital Twin Trial of a cognitive decline-slowing AD drug treatment. Compared to a standard RCT analysis, our DTT analysis empirically increased power and reduced required sample size in simulations of AD trials constrained by the inclusion criteria and duration of lecanemab’s Phase 3 trial CLARITY AD.^29,30^ Our DTT simulations also demonstrated Type 1 error control. All simulations used data on real ADNI participants.

In Study 1, there were no statistically significant differences on key demographic and clinical factors between the Recruited Cohort and the External Cohort. While the Recruited Cohort’s model-estimated decline trend appears slightly steeper in Fig. 2 compared to the External Cohort’s trend, the slopes were not significantly different. This similarity is not surprising because both cohorts were pulled from ADNI and satisfy the same CLARITY AD criteria. However, subjects in the Recruited Cohort were from different real-world ADNI sites than External Cohort subjects. This geographical difference could explain the visual difference in decline trends.

We evaluated our DT model’s accuracy at predicting 18-month CDRSBΔbl scores of subjects in our Recruited Cohort using Twins identified in our External Cohort. The DPP that contained core clinical and demographic variables yielded prediction accuracies that were not significantly different from accuracies of the maximal DPP that also included BMI and education. The minimal DPP that excluded baseline cognitive categories and MMSE yielded worse predictions than the core and maximal DPPs. The core and maximal DPPs both yielded better predictions than ones made using randomly selected Twins. DPP set 1 and *n*_*Twins*_ = 20 balanced prediction accuracy and model parsimony. We used this combination in Studies 2 and 3.

In Study 2, our DTT required approximately 1,855 recruited subjects to detect a 25% decline-slowing drug effect at 0.9 power. To detect the same effect, our sRCT required approximately 2,170 recruits. This DTT sample size is approximately 14.5% smaller than the sRCT’s. Depending on a trial’s cost per participant and the recruitment rate at each data collection site, a 14.5% sample size reduction could reduce a trial’s total run time by several months and save millions of dollars in expenditures.^3,49^

Our DTT power gain is achieved by leveraging our model’s predictions within PPI, which adjusts the trial’s outcome metric to reduce its variance in Drug and Placebo groups. This variance reduction makes drug effect estimates more precise, which increases statistical power and reduces the number of recruits required for the trial to detect a subtle drug effect.

In Study 3, the DTT and sRCT Type 1 error probabilities were not significantly different across sample sizes, and no probability was significantly different from 0.05. This result indicates empirically that our DTT controls Type 1 error in a realistic trial parameter space.

While our DT model made modestly accurate predictions in Study 1, the accuracy was sufficient to produce the observed power gain. Future model versions will improve Twin identification, for example by optimizing weightings of variables in the similarity calculation and by testing different similarity estimators (e.g., cosine).^50^ This optimization will improve prediction accuracy and further reduce required sample sizes for DTTs.

Our DTT approach addresses a key limitation of unadjusted external cohort trial analyses, where an external cohort dissimilar to a trial’s can inflate error risk. Bayesian dynamic borrowing methods can address this issue by weighting the contribution of external data based on their similarity to trial data, but the requirement for careful specification of model assumptions can complicate regulatory acceptance.^14–16^

Propensity scoring techniques can be used to account for differences between trial and external cohorts by weighting or selecting external subjects based on their similarity to the average characteristics of the trial cohort.^51,52^ However, unmeasured confounders in propensity scoring can still introduce bias, which increases error risk.

Our framework assumes that any systematic bias from unmeasured confounders or inaccurate predictions is equally distributed to Drug and Placebo groups. In the group comparison step, PPI cancels out this bias and thus mitigates inflation of Type 1 and 2 error risk.^17^

While predictive DT models have been leveraged in AD trials to increase power, those models have relied on more opaque machine learning techniques to predict patient outcomes.^17,21,23,26,27^ Clinicians have hesitated to trust such techniques.^53^ On the other hand, our DT model is highly interpretable and straightforward to implement: a patient is matched to medically similar real individuals at baseline from existing AD databases; the Twins’ observed outcome trajectories are averaged to predict the patient’s disease progression.^5^

Also, our DT model preserves inter-participant variability within the External Cohort, a key advantage over highly parametrized black box models that only capture partial covariance among prognostic factors and subsequent disease progression. Moreover, in our framework, if a trial’s efficacy metric is available in the External Cohort at the same follow up times as the trial’s (e.g., within the same visit window), then no model training or interpolation is required. These characteristics distinguish our technique from commercially available and more complex deep learning techniques.^54^

The Type 1 error and power analyses in this study all used our recently proposed simulation technique that combines real AD patient data with statistical models of disease progression.^31,32,34^ Parametric simulations may not adequately capture AD heterogeneity or key trial features, but our simulation technique does so by sampling real patient data. Our technique lets us evaluate how trial-specific design parameters (e.g., duration and inclusion criteria) affect a trial’s ability to detect posited drug effects while accounting for realistic AD heterogeneity.

This study and our DT technique have some limitations. First, prediction-leveraged analyses enhance the contrast between treatment groups, so a group imbalance in fast or slow decliners might increase Type 1 and 2 error risk compared to a standard analysis. However, by running 3,000 simulations of our DTT and sRCT analyses, the likelihood and effect of such imbalances on power and Type 1 error risk are factored implicitly into our results.

In Study 1 we did not cross validate our prediction model because of the limited data available in ADNI and the requirement that selected subjects satisfy the posited CLARITY AD inclusion criteria.^29,30^ However, we avoided data leakage in our analyses by first splitting our ADNI pool roughly in half by geographical site into a Recruited Cohort and an External Cohort. We predicted Recruited Cohort subjects’ decline using Twins selected in the independent External Cohort.

ADNI is also a North American study of mostly white, highly educated males.^28,55,56^ Future work will run 10-fold cross validation and evaluate fully out-of-sample prediction accuracy using a more diverse pooled dataset comprising patients from ADNI and other large AD datasets from the Australian Imaging, Biomarkers and Lifestyle Study of Aging (AIBL), the National Alzheimer’s Coordinating Center (NACC), and other organizations.^57,58^

The External Cohort in our method must satisfy the same inclusion criteria as the trial’s Recruited Cohort. However, unmeasured factors such as evolving AD diagnostic criteria could enhance differences between a modern Recruited Cohort and External Cohort drawn from years-old retrospective datasets, despite both satisfying the same criteria.^10–13^ Geography and socioeconomic status could also produce differences between cohorts.^59,60^ While PPI distributes bias equally to treatment groups in a trial, careful consideration must still be given to selecting the External Cohort. Future work will examine DTT power and Type 1 error sensitivity to dissimilarity between Recruited and External Cohorts, i.e., scenarios where the conditional exchangeability assumption described in section 2.4.2 is violated.

ADNI subjects receive standard of care, not placebo. Future work will therefore use External Cohorts constructed using control group data from past trials, such as those pooled and shared openly in the Critical Path for Alzheimer’s Disease (CPAD) dataset.^61^ Privately shared control group data from past trials within pharmaceutical companies can also be used. Future work would benefit from testing our method in retrospective control and real drug group data from past trials. The open sharing of trial data by pharmaceutical companies could accelerate this work.

Prospective DTT validation in strictly regulated ongoing trials can help identify and correct model biases. Such validation of our interpretable model could foster clinician trust and smooth the model’s implementation in the clinic, where it could be used to make individualized prognoses and evaluate treatment effects in individual patients.^53^

Our study used linear modeling in our interpolation steps despite cognitive decline in AD having a nonlinear trend over the full disease timeline.^62^ However, linear assumptions are reasonable over the 18-month period of a trial. Future work could nonetheless implement nonlinear models to more comprehensively map cognitive decline trajectories.

The ratio of subjects in intervention and control groups can also be optimized based on model prediction accuracy. In PPI, more accurate predictions imply fewer subjects are needed in the control group than the intervention group.^17^ However, the sample size reduction we demonstrated here could already help accelerate AD trials. Our prediction model will also support precision medicine by evaluating drug effects in individual trial participants.

## Data Availability

All data used in preparation of this article were obtained from the Alzheimer's Disease Neuroimaging Initiative (ADNI) database (https://adni.loni.usc.edu).

## 6. Acknowledgments

Data collection and sharing for the Alzheimer’s Disease Neuroimaging Initiative (ADNI) is funded by the National Institute on Aging (National Institutes of Health Grant U19AG024904). The grantee organization is the Northern California Institute for Research and Education. In the past, ADNI has also received funding from the National Institute of Biomedical Imaging and Bioengineering, the Canadian Institutes of Health Research, and private sector contributions through the Foundation for the National Institutes of Health (FNIH) including generous contributions from the following: AbbVie, Alzheimer’s Association; Alzheimer’s Drug Discovery Foundation; Araclon Biotech; BioClinica, Inc.; Biogen; Bristol-Myers Squibb Company; CereSpir, Inc.; Cogstate; Eisai Inc.; Elan Pharmaceuticals, Inc.; Eli Lilly and Company; EuroImmun; F. Hoffmann-La Roche Ltd and its affiliated company Genentech, Inc.; Fujirebio; GE Healthcare; IXICO Ltd.; Janssen Alzheimer Immunotherapy Research & Development, LLC.; Johnson & Johnson Pharmaceutical Research & Development LLC.; Lumosity; Lundbeck; Merck & Co., Inc.; Meso Scale Diagnostics, LLC.; NeuroRx Research; Neurotrack Technologies; Novartis Pharmaceuticals Corporation; Pfizer Inc.; Piramal Imaging; Servier; Takeda Pharmaceutical Company; and Transition Therapeutics.

## 7. Conflict of Interest Statement

All authors declare no conflicts of interest.

## 8. Funding sources

This work was supported by McGill University, the Famille Louise & André Charron, Brain Canada, the Canadian Institutes of Health Research (Daniel Andrews: Frederick Banting and Charles Best Canada Graduate Scholarships Doctoral Award; Prof. D. Louis Collins: CIHR Project Grant FRN 165921), and the Natural Sciences and Engineering Research Council of Canada (Prof. D. Louis Collins: NSERC Discovery Grant RGPIN-2015-03633).

## 9. Consent statement

ADNI obtained ethics approval from all participating institutions, and all participants provided informed consent.

## Notes

### Competing Interest Statement

The authors have declared no competing interest.

### Author Declarations

All data used in preparation of this article were obtained from the Alzheimer's Disease Neuroimaging Initiative (ADNI) database (https://adni.loni.usc.edu) following the approval by ADNI of our data access request. ADNI obtained ethics approval from all participating institutions, and all ADNI participants provided informed consent.

